# A Data-Driven Approach for Linking Epileptic Networks and Cognitive Profiles Using Stereo-EEG

**DOI:** 10.64898/2026.04.29.26352098

**Authors:** Parveen Sagar, Emily Cockle, Thanomporn Wittayacharoenpong, Alissandra McIlroy, Jacob Bunyamin, Joshua Laing, Matthew Gutman, Martin Hunn, Patrick Kwan, Terence J. O’Brien, Matthew Hudson, Genevieve Rayner, Andrew Neal

## Abstract

**Objective:** Neuropsychological assessment plays an important role in localizing epileptogenic regions during presurgical evaluation. However, its diagnostic potential is constrained by reliance on syndromic models of epilepsy. A network-grounded approach may provide higher resolution structure–function relationships, yet in vivo evidence linking epileptogenic networks to cognitive deficits remains limited. Here, we developed a network-based framework to test the hypothesis that patterns of sublobar epileptogenicity shape distinct cognitive profiles.

**Methods:** Retrospective cohort study of 42 drug-resistant focal epilepsy patients undergoing stereo-EEG (SEEG) and pre-implantation neuropsychological assessment (16 indices). Epileptic networks were quantified using a composite SEEG-derived epileptogenicity metric (‘EzPz’ score) providing a continuous measure of regional epileptogenicity. Sublobar EzPz values and neuropsychological z-scores were analyzed by Pearson correlations, PCA, and hierarchical clustering to derive network subtypes and domain-specific cognitive associations.

**Results:** Mean age 34.9 years and 79% MRI-negative. Significant negative pairwise correlations were seen: dominant temporal with language, non-dominant mesiotemporal with visual memory; and non-dominant frontal with attention and visuospatial. Exploratory PCA/clustering identified nine network configurations with associated cognitive profiles. For example, dominant mesiolateral temporal configuration (86% MRI-negative): naming impairment with preserved verbal memory; non-dominant frontotemporal: severe executive and visual memory impairment; bimesiolateral temporal: severe language deficits with executive and visuoconstructional impairment.

**Interpretation:** Each of our nine network configurations were associated with a cognitive profile shaped by sublobar epileptogenic distribution, lateralisation, and network size. These findings support a shift from syndromic to network-based interpretation of neuropsychological data. Our framework may enhance the diagnostic potential of neuropsychological assessment in SEEG hypothesis generation and surgical planning.

## Introduction

Neuropsychological assessment is a cornerstone of presurgical evaluation in epilepsy, but its diagnostic utility is constrained by longstanding localizationist models of brain function, which have not fully kept pace with contemporary network-based understandings.^1^ Historically, neuropsychological tools have developed alongside epilepsy taxonomies, which classically classify focal epilepsies into syndromes such as temporal and frontal lobe epilepsies (TLE, and FLE).^1^ Cognitive signatures have been accordingly formulated, presupposing lobe-specific patterns of impairment.^1,2^

In contrast, the brain is now widely understood as an interconnected network.^3,4^ Crucially, both the physiological processes supporting cognition and pathological processes that drive epilepsy are recognized as properties of large-scale brain networks.^4–6^

That epilepsy is a network disorder is well established.^7^ Existing biomarkers of epileptogenicity provide quantitative evidence of epilepsy’s distributed architecture.^8^ These findings demonstrate that syndromic labels such as TLE and FLE represent pragmatic simplifications of complex networks. Each syndrome encompasses distinct subtypes—such as mesial or polar variants—each with its characteristic topology and anatomy. This underlying diversity raises concerns about relying on broad syndromic classifications to guide diagnostic neuropsychology.

For example, TLE is classically associated with memory and language dysfunction, while FLE with attentional and executive dysfunction.^9^ However, cognitive impairments are increasingly recognized to be heterogeneous within epilepsy syndromes.^10^ In response, recent work has sought to characterise patterns of impairment, or ‘cognitive phenotypes’ within epilepsy syndromes, and to identify their structural correlates^11,12^, that have been shown to extend beyond the namesake lobe.^13–15^ The IC-CoDE framework was recently developed to standardize the identification of these phenotypes.^16^ While these phenotypes and their associations with widespread abnormalities lend support to a network understanding of cognitive dysfunction, the IC-Code framework remains syndromically anchored.

The diagnostic utility of neuropsychology to localise regions of epileptogenicity can likely be enhanced by shifting from a syndrome-based to a network-led framework.^6^ However, in vivo evidence linking epileptogenic networks to cognitive dysfunction is scant, in part due to the spatial and temporal constraints of available imaging and electrophysiological methods.

In this proof-of-concept study, we describe a framework for characterising the relationship between epileptic networks and cognitive profiles. Few methods currently allow for systematic subtyping of epileptic networks in vivo or for linking these networks to cognitive outcomes. We addressed this gap by mapping epileptogenicity at the sublobar level using a SEEG-derived metric that integrates both ictal and interictal features to quantify epileptogenicity. We used this metric to characterize the epileptic network spanning the epileptogenic zone (EZ), and the propagation zone (PZ). The PZ inclusion rests on the premise that cognitive impairment may result from network dysfunction in regions beyond the EZ, consistent with the concept of a “functional deficit zone”.^17^ Numerous structural and functional imaging studies demonstrate network abnormalities beyond the EZ that correlate with the severity and nature of cognitive impairment.^15,18,19^ We hypothesised that different sublobar distributions of epileptogenicity would be associated with unique cognitive impairment patterns.

This study pursued two aims: (1) to identify distinct epileptic networks at the sublobar level using a quantitative marker of epileptogenicity; and (2) to determine whether these networks are associated with cognitive profiles that reflect the underlying distribution of epileptogenicity.

## Materials and methods

### Study design and participants

Consecutive adults with DRFE who underwent SEEG at the Alfred Hospital, Melbourne, Australia from 2018 to 2023 were identified. The number and location of SEEG electrodes was guided by the epileptic network hypotheses following an evaluation of non-invasive data and discussion in a multidisciplinary team meeting. As part of their pre-SEEG workup, all patients had a minimum standardised battery of neuropsychological testing performed (Table 1 displays the tests utilized.^20–27^ Performance was normalised to population data using z-scores).

**Table 1:**
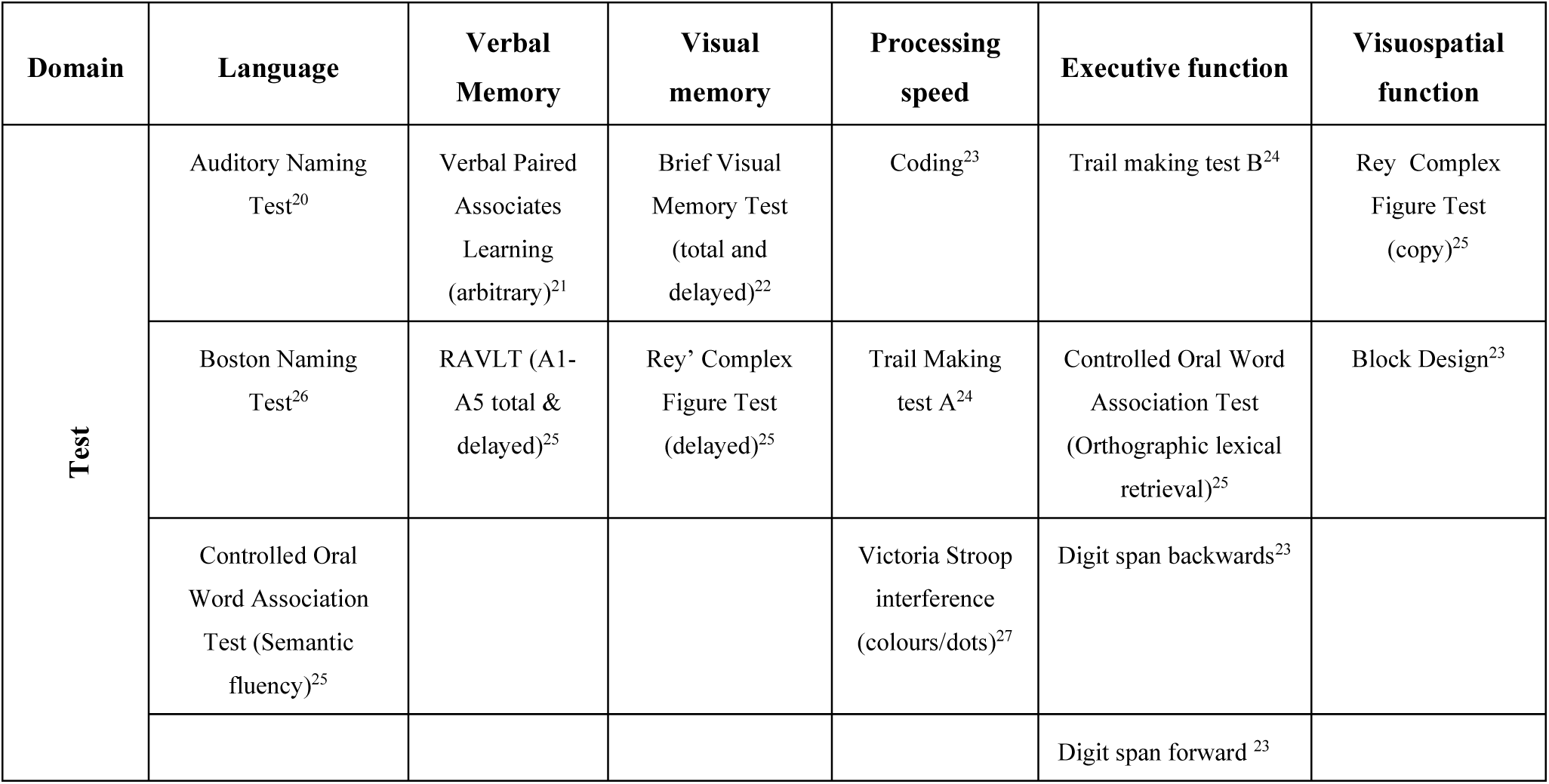
Testing battery administered as part of pre-SEEG workup, organized by the targeted cognitive domain.

| Domain | Language | Verbal Memory | Visual memory | Processing speed | Executive function | Visuospatial function |
| --- | --- | --- | --- | --- | --- | --- |
| Test | Auditory Naming Test<sup>20</sup> | Verbal Paired Associates Learning (arbitrary)<sup>21</sup> | Brief Visual Memory Test (total and delayed)<sup>22</sup> | Coding<sup>23</sup> | Trail making test B<sup>24</sup> | Rey Complex Figure Test (copy)<sup>25</sup> |
|  | Boston Naming Test<sup>26</sup> | RAVLT (A1-A5 total & delayed)<sup>25</sup> | Rey' Complex Figure Test (delayed)<sup>25</sup> | Trail Making test A<sup>24</sup> | Controlled Oral Word Association Test (Orthographic lexical retrieval)<sup>25</sup> | Block Design<sup>23</sup> |
|  | Controlled Oral Word Association Test (Semantic fluency)<sup>25</sup> |  |  | Victoria Stroop interference (colours/dots)<sup>27</sup> | Digit span backwards<sup>23</sup> |  |
|  |  |  |  |  | Digit span forward <sup>23</sup> |  |

**Table 2:** Baseline clinical and demographic characteristics of the SEEG cohort (n = 42).

| Characteristics for n=42 | Value |
| --- | --- |
| Age at SEEG (years) | 34.9 ± 9.3 |
| Sex - Female (n, %) | 24 (57%) |
| Epilepsy duration (years) | 19.4 ± 11.7 |
| Number of ASMs at SEEG | 3.0 ± 0.9 |
| MRI: Lesional (n, %) | 9 (21%) |
| PET Focal hypometabolism (n, %) | 33 (79%) |
| Language dominance (fMRI and cortical stimulation) | Left: 31 (74%) |
|  | Other: 11 (26%) |
| Implant laterality | Bilateral: 21 (50%) |
|  | Left: 14 (33%) |
|  | Right: 7 (17%) |
| Number of electrodes implanted | 14.0 ± 2.9 |

Out of 52 patients, 42 met inclusion criteria - adult DRFE patients undergoing SEEG with complete pre-implantation neuropsychological data. Patients were excluded if (i) their neuropsychological data was missing or deemed unreliable by the neuropsychologist (n=3), (ii) if no seizures were captured during their SEEG admission (n=1), and (iii) if they had prior resections (n=6) as the baseline deficits may be driven by anatomical resections rather than epileptogenic networks. Study size reflects proof-of-concept design using all available complete cases.

SEEG recordings were performed using multi-contact intracerebral electrodes, ranging from 8 to 18 contacts (manufactured by Dixi Medical).

Patients underwent video-SEEG (Compumedics, Australia), with signals recorded with a bandpass filter between 0.5Hz and 1000Hz and with a sampling rate of 2000Hz. Patients underwent a planned antiseizure medication withdrawal with the aim of capturing habitual seizures.

The study was approved by the Human Research Ethics Committee of Alfred Health, Melbourne, Australia.

### Standardising brain region labels across patients

Post-implantation computerized tomography was obtained and co-registered to pre-operative MRI to determine sublobar contact localisation. We categorised each contact according to 14 pre-defined regions adapted from previously published work (Supplementary Table 1).^28^

### SEEG-derived epileptogenicity metric

To quantify epileptogenic involvement across the epileptic network, we required a metric capable of representing regional epileptogenic burden as a continuous gradient. Because no single existing biomarker simultaneously captures both seizure onset and propagation dynamics across diverse seizure onset patterns, we constructed a pragmatic composite SEEG-derived metric termed the Epileptogenic Zone–Propagation Zone (EzPz) score. The aim of this metric was not to establish a new validated biomarker, but to provide a quantitative representation of epileptogenic burden suitable for network-level analysis in this exploratory framework. Several established biomarkers quantify different aspects of epileptogenicity, but were not suitable for our aims. Interictal biomarkers, such as spike and HFO rates, are broadly applicable but do not consistently localize the EZ.^29,30^ While emerging composite approaches like gamma-spikes or spike-ripples^31,32^ show promise in improving specificity, their utility remains under evaluation.

Ictal biomarkers generally outperform interictal markers at detecting the EZ. However, measures such as the “epileptogenic fingerprint”^33^ and “chirp”^34^ are tuned specifically to detect the seizure source and do not account for the PZ. The Epileptogenicity Index (EI) ^35^ can provide a graded measure across the network, but it is designed primarily for seizures with low-voltage fast activity (LVFA) onset. This excludes approximately 20% of seizures beginning with alternative electrographic patterns.^36^

To enable our network-level analysis and ensure applicability across diverse seizure onset patterns, we constructed an SEEG-based metric integrating ictal and interictal features. The metric’s derivation was guided by two primary principles: (i) that the epileptic network must encompass regions of both seizure onset and propagation; and (ii) that greater regional epileptogenicity is reflected by both earlier ictal recruitment and higher interictal discharge density. By combining these features into a single contact-wise measure, the EzPz score provides a pragmatic representation of distributed epileptogenicity suitable for examining relationships between epileptic network organization and cognitive phenotypes.

Our epileptogenicity metric was developed from dataset of 1,236 SEEG contacts from 13 patients who were rendered seizure-free for at least six months following radiofrequency thermocoagulation (RF-THC). Among these, 135 contacts had been thermocoagulated, and 1,101 had not. Candidate predictors included spike rate, HFO rate, spike x HFO rate, and an ‘ictal score’ (described below). Using univariate analyses and logistic regression, we systematically evaluated combinations of these variables to identify the model with the strongest predictive value for classifying RF-THC (as a surrogate for being part of EZ) contacts.

The final model, selected using the Bayesian Information Criterion, retained spike rate and ictal score as independent predictors. Model discrimination in the development cohort was then evaluated across five repeated subsamples, yielding stable results across all iterations (AUC = 0.97; Youden index = 0.721). The final logistic regression equation was:

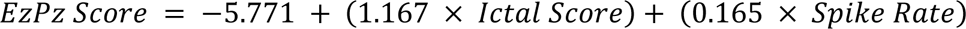

The optimal classification threshold was identified using the Youden index (−0.8876), corresponding to a sensitivity of 0.833 and specificity of 0.88 within the development dataset. Scores above this threshold were interpreted as reflecting greater likelihood of involvement in the EZ, while lower scores were interpreted as PZ, forming a continuous gradient of epileptogenicity.

The distribution of EzPz scores across thermocoagulated and non-thermocoagulated contacts in both the development cohort (n = 13) and the full clinical cohort (n = 52) is shown in Supplementary Fig. 2A and 2B.

### Deriving Quantitative Features for EzPz Score

For every patient, a 20min interictal sleep segment was analysed using Delphos, an automated detector of oscillations and spikes, to quantify HFO, and spike-rate and HFO cross rate (square root of HFO x spikes) for every implanted grey matter channel. Artefactual channels were excluded via visual inspection.

For ‘ictal scoring’, each patient’s habitual electroclinical seizures were visually inspected. Ictal scoring was performed by an experienced rater blinded to the patients’ pre-implantation neuropsychological profiles. An ictal score was assigned to each channel based on the timing of its involvement in the ictal discharge: channels involved within the first second were scored as 5; those involved between 1–3 seconds, as 3; between 3–10 seconds, as 1; and beyond 10 seconds, as 0. This non-linear weighting scheme reflects the assumption that earlier involvement carries greater epileptogenic significance. The final ictal score for each channel was calculated as the arithmetic mean across all seizures. To ensure the ictal score reflected individual the ictal network dynamics of each patient, we employed a representative sampling strategy. Sampling of each habitual electroclinical seizure was performed until electrographic stereotypy were confirmed; additional seizures exhibiting identical spatiotemporal patterns to those already scored were excluded to avoid over-representing redundant ictal events.

### Statistical analysis

Statistical analysis was informed by the methodology described by Bonini et al.^37^ Two matrices were derived: (1) *neuropsychologic test* x *patients (z-scores)*, and (2) *brain regions* x *patients* (EzPz scores). Ordinal versions were created to emphasize clinical categories (EZ/PZ/no involvement) and stabilize PCA with sparse SEEG data For the EzPz matrix, values were binned as follows: 2 = EzPz > –0.86 (presumed EZ), 1 = EzPz ≤ –0.86 (presumed PZ), and 0 = visually scored 0 (no ictal involvement). For neuropsychological scores, bins were: > –1.5 = 0 (no impairment), –1.5 to –2 = 1 (mild), –2 to –2.5 = 2 (moderate), and < –2.5 = 3 (severe). Continuous matrices were retained for correlation analyses.

As an exploratory first step, we assessed whether cognitive performance was associated with epileptogenicity in individual brain regions using pairwise Pearson correlations. We hypothesised that higher epileptogenicity in specific regions would correlate with domain-specific deficits. This analysis was performed using SciPy (Python); *p* < 0.05 was considered significant.

Next, we aimed to identify groups of patients with shared patterns of epileptogenicity. We hypothesized that patients would cluster into distinct epileptic network groups based on the distribution of epileptogenicity across brain regions. PCA was applied to the ordinal EzPz matrix using scikit-learn, reducing dimensionality by converting correlated variables (brain regions) into a smaller set of uncorrelated components. To address missing data due to variable implantation, unsampled regions were assumed uninvolved and assigned EzPz = –6 (below the first percentile of observed scores), reflecting typical SEEG targeting strategy.

The first six components were retained, explaining 72% of the variance. Hierarchical clustering using Ward-linkage on the PCA-derived coordinates was subsequently performed, using SciPy to identify patient clusters based on network structure. The number of clusters was selected based on the silhouette score and informed by clinical knowledge to ensure the most meaningful partitioning of the data.

Patients were grouped into relatively homogenous epileptic network clusters. For each cluster, we characterised the network by calculating mean EzPz scores across brain regions (epileptogenic burden) and identified statistically distinctive regions using the V-test statistic applied to the ordinal dataset. The V-test measures the deviation of a region’s mean score within a cluster from the overall cohort mean. Each cluster’s cognitive profile was then characterised by computing the median ordinal neuropsychological scores of its members. Finally, we qualitatively examined the anatomical and cognitive patterns of each cluster to identify network configurations and interpret structure–function relationships.

## Results

### General characteristics

Following SEEG, 13 patients were seizure-free at six months following RF-THC. Thirty-two patients had been offered resective surgery or LITT, of whom 16 underwent the procedure with available outcome data. Among these, seven patients (43.8%) achieved Engel class I outcome at 12 months post-surgery or LITT. The duration of follow-up among patients with Engel I at last available follow-up (n = 9) ranged from 6.4 to 40.3 months (median: 15.2 months). Taken together, 22 of 29 patients (75.9%) who received definitive treatment (RF-THC, surgery, or LITT) were seizure-free at follow-up (defined as 6 months for RF-THC, and 12 months or last follow-up for surgery/LITT).

A total of 650 seizures were recorded during SEEG exploration. A median of 7.3 seizures were recorded per patient. A total of 95 representative seizures contributed to the ictal score in the final analysis, with a mean of 2.5, and median of 2 (range: 1-8) seizures analysed per patient.

### Regional Epileptogenicity Correlates with Domain-Specific Cognitive Impairment

Fig. 1 depicts a correlation matrix illustrating the relationship between neuropsychological test performance and epileptogenicity across brain regions. Negative correlations indicate that higher epileptogenicity is associated with poorer cognitive performance. Supplementary Fig. 1 presents the correlation matrix with annotated correlation coefficients. In the text below, only the strongest negative correlations within each cognitive domain are highlighted.

**Figure 1:**
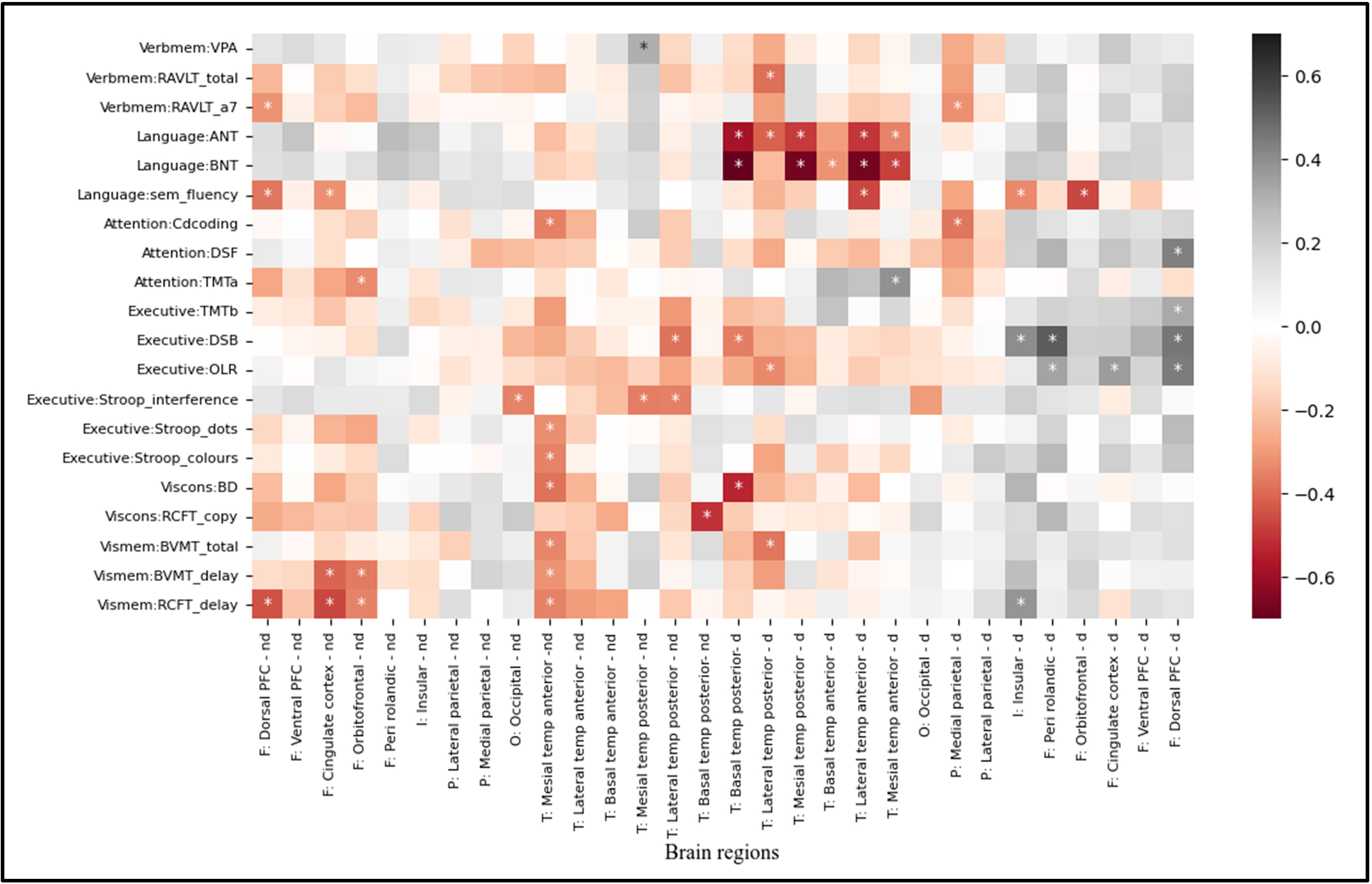
Correlation matrix between epileptogenicity of brain regions and performance on neuropsychological indices. Pearson correlation matrix depicting the relationship between epileptogenicity of brain regions versus performance on individual NP measures. Significance represented by asterisk: p<0.05. Colour scale from grey to deep red, reflects the change in correlation coefficient: red=negative correlation – higher epileptogenicity correlates with greater impairment, and grey=positive correlation – the converse. NP tests axis labelled so: ‘Domain: test’. Domains: verbmem=verbal memory, viscons= visuoconstructional, vismem=visual memory. Tests: VPA=verbal paired associates, RAVLT_total=Rey Auditory Verbal Learning Test, ANT=auditory naming test, BNT=Boston naming test, Sem_fluency=semantic fluency, DSF=digit span forward, DSB=digit span backwards, OLR=orthographic lexical retrieval, BD=block design, RCFT=Rey complex figure test, BVMT=brief visual memory test. Brain regions are labelled so: ‘Lobe: brain region-dominance’. Lobes: F=frontal, I=insular, P=parietal, T=temporal, O=occipital. Dominance: nd=non-dominant, d=dominant. * p<.05.

Significant and strongly negative correlations are observed between dominant temporal lobe epileptogenicity and measures of language (r=-0.71). A weak, non-significant negative correlation was observed between dominant mesial temporal regions and verbal memory measures (r=-0.16). The non-dominant mesial temporal region also shows significant but weakly negative correlations with visual memory (r= –0.35), visuoconstructional (r= –0.38), and executive function (r= –0.32) measures. Non-dominant frontal regions show negative correlations with visual memory measures (r= −0.34), while dominant frontal regions display relatively positive correlations, suggesting preserved function. These findings suggest region-specific epileptogenicity is linked to domain-specific cognitive deficits.

### Principal Component Analysis and hierarchical clustering

Based on PCA-derived coordinates, patients were hierarchically clustered into nine electroclinically meaningful groups. The silhouette score was used to assess clustering structure across candidate solutions. The nine-cluster solution produced the highest silhouette score (0.30), indicating modest but positive cluster separation. This solution (Fig. 2) was retained as it aligned with clinically recognised SEEG network configurations and preserved distinctions that were lost when the dendrogram was cut at higher levels.

**Figure 2:**
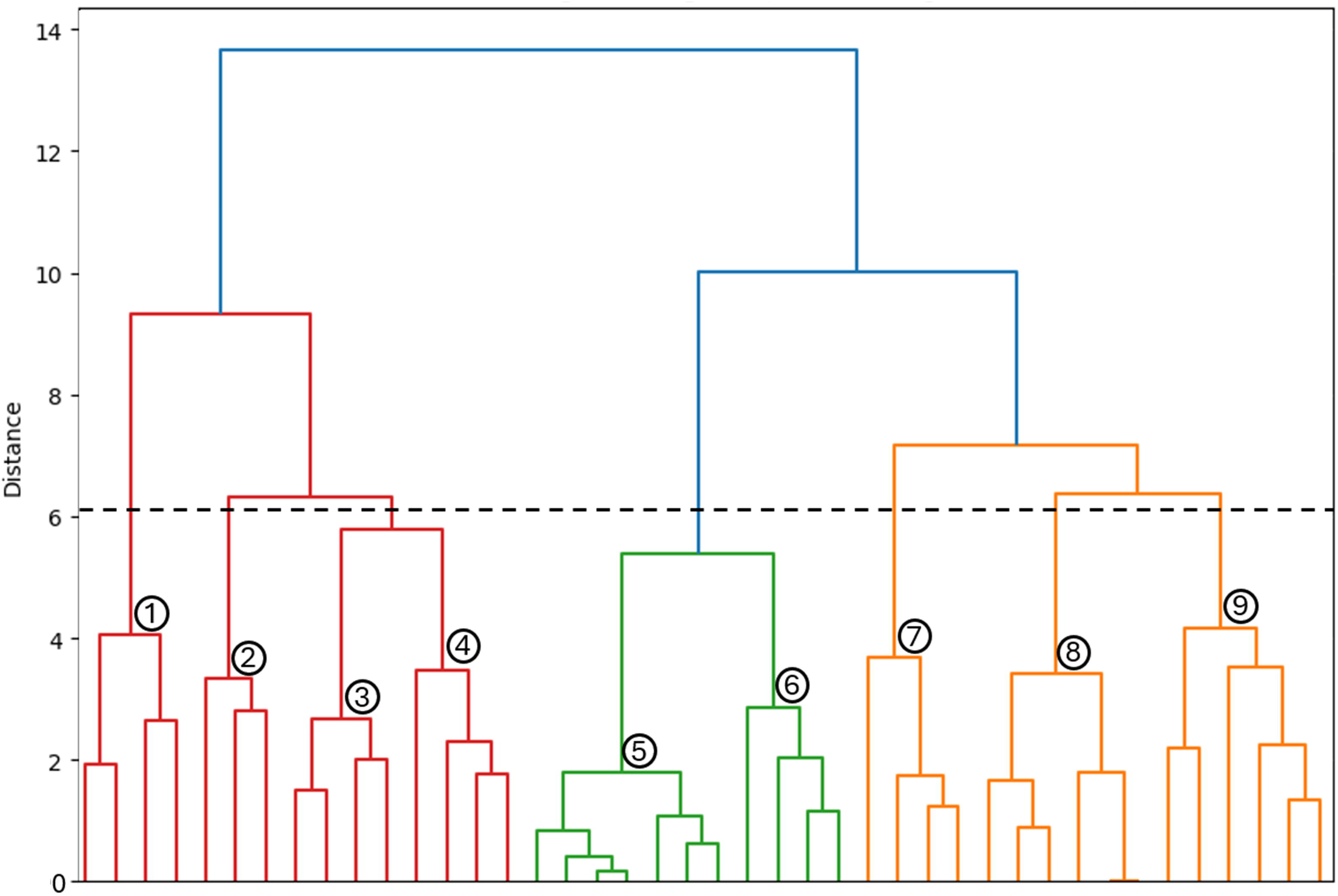
Ward’s Hierarchical Clustering on PCA-Transformed Epileptogenicity (EzPz) Scores. Hierarchical clustering dendrogram using Ward’s linkage on PCA-transformed EzPz data. Nine clusters were selected by cutting the dendrogram at the threshold indicated by the dashed line.

### Characterisation of epileptic networks clusters

The epileptogenic profile of each cluster was identified through region-wise V-test analysis of EzPz scores, with statistically distinctive network patterns emerging (Fig. 3A), such as mesiotemporal, bimesiolateral temporal, and frontotemporal networks. Table 3 summarizes demographic and clinical features across clusters. Qualitatively, age, sex, number of ASMs, and structural imaging findings were broadly comparable, although differences in premorbid cognitive function (measured by Test of Premorbid Function; TOPF), and network size were noted in select clusters. Network size was defined as the number of brain regions within a cluster that were involved in the EZ or PZ (i.e. EZPZ score > 0).

**Figure 3:**
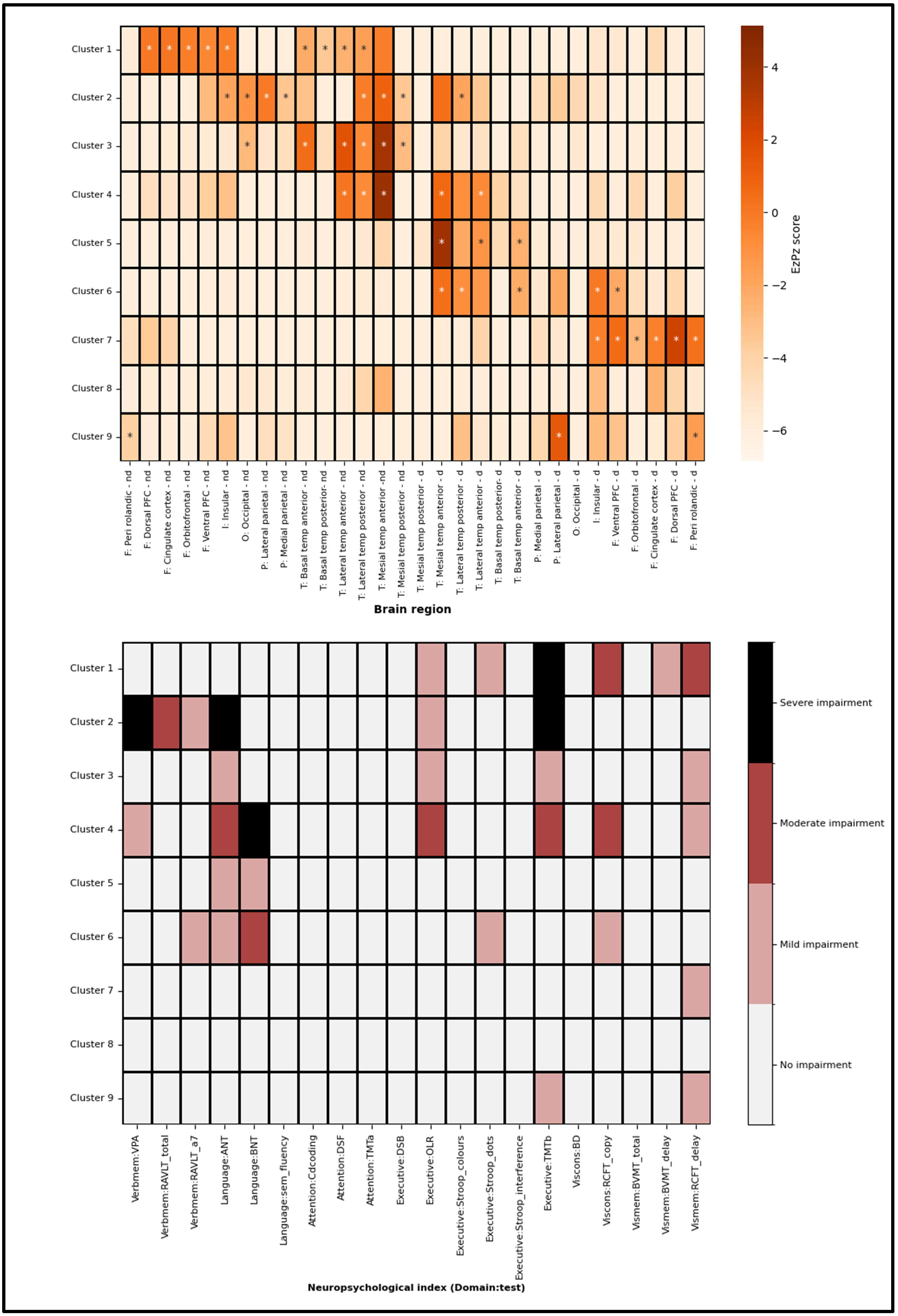
Epileptogenic Network and Neuropsychological Profiles Across Clusters A. Mean EzPz scores per brain region across clusters, representing the distribution of epileptogenicity within each network. Warmer colours indicate higher epileptogenicity. Asterisks (*) mark regions that meet both statistical and clinical relevance thresholds (V-test > 1.65 and EZPZ > −4.), identifying regions that are both distinctive for the cluster and carry significant epileptogenic burden. **B:** Median neuropsychological impairment levels per cluster across test indices. Impairment severity is categorized as: No impairment (0–1), Mild (1–2), Moderate (2–3), and Severe (>3), as defined in the accompanying colour key. See Figure 1 caption for full neuropsychological test name abbreviations.

**Table 3:** Demographic and Clinical Characteristics of Epileptic Network Clusters.

| Cluster # (n) and name | Sex (female) | Age (mean) | Lang. dom. (left) | MRI negative | FDG-PET negative | No. ASM at time of SEEG | Epilepsy duration (years) | TOPF<sup>2</sup> (mean z-score) | Unique brain regions involved in network (total and mean per person)<sup>3</sup> |
| --- | --- | --- | --- | --- | --- | --- | --- | --- | --- |
| 1 (n=4)<br>**non-dominant frontotemporal** | 0 | 38+/-13.6 | 4 | 4 | 3 | 3+/-0.8 | 23.26 | -0.2 | total: 15<br>mean: 4.5 |
| 2 (n=3)<br>**bilateral TPO<sup>1</sup>** | 2 | 42.3+/-9.0 | 2 | 2 | 0 | 2.3+/-1.2 | 21.27 | -0.8 | total: 16<br>mean: 5 |
| 3 (n=4)<br>**non-dominant mesiolateral temporal** | 3 | 35.5+/-8.0 | 4 | 2 | 0 | 3.2+/-1.3 | 19.23 | -0.6 | total: 23<br>mean: 3.5 |
| 4 (n=4)<br>**bimesiolateral temporal** | 3 | 42.6+/-11.3 | 4 | 3 | 1 | 3+/-1.0 | 27.84 | -1.5 | total: 17<br>mean: 4.3 |
| 5 (n=7)<br>**dominant mesiolateral temporal** | 6 | 38.4+/-6.9 | 6 | 6 | 1 | 2.9+/-0.7 | 14.48 | -0.6 | total: 19<br>mean: 1.6 |
| 6 (n=4)<br>**dominant frontotemporal** | 2 | 38.5+/-2.4 | 4 | 3 | 0 | 3.3+/-1.0 | 12.92 | -0.1 | total: 15<br>mean: 2.8 |
| 7 (n=4)<br>**dominant frontal** | 3 | 32.2+/-11.1 | 2 | 3 | 1 | 2.8+/-1.3 | 17.81 | 0 | total: 14<br>mean: 4.8 |
| 8 (n=6)<br>**no clear pattern** | 2 | 34.3+/-11.3 | 6 | 4 | 2 | 2.7+/-1.0 | 19.79 | -0.2 | total: 23<br>mean: 1.3 |
| 9 (n=6)<br>**dominant parietofrontal** | 3 | 34.6+/-11.5 | 5 | 6 | 1 | 3.3+/-0.8 | 20.83 | -0.2 | total: 19<br>mean: 2.3 |

<sup>1</sup>temporoparietooccipital, <sup>2</sup>test of premorbid function, <sup>3</sup>network size = number of regions with non-zero EzPz scores (EZ or PZ involvement), reported as total and mean per patient.

Figures 3 and 4 illustrate the epileptogenic and cognitive profiles of each cluster. Fig. 3A shows sublobar patterns of epileptogenicity, while Fig. 3B displays corresponding domain-level impairments. Fig. 4 visualises each cluster’s network using a cortical atlas, with adjacent tables summarising the cognitive patterns. Together, these figures show that each cluster is characterised by a distinct combination of epileptogenicity and displays a unique set of associated cognitive deficits.

**Figure 4:**
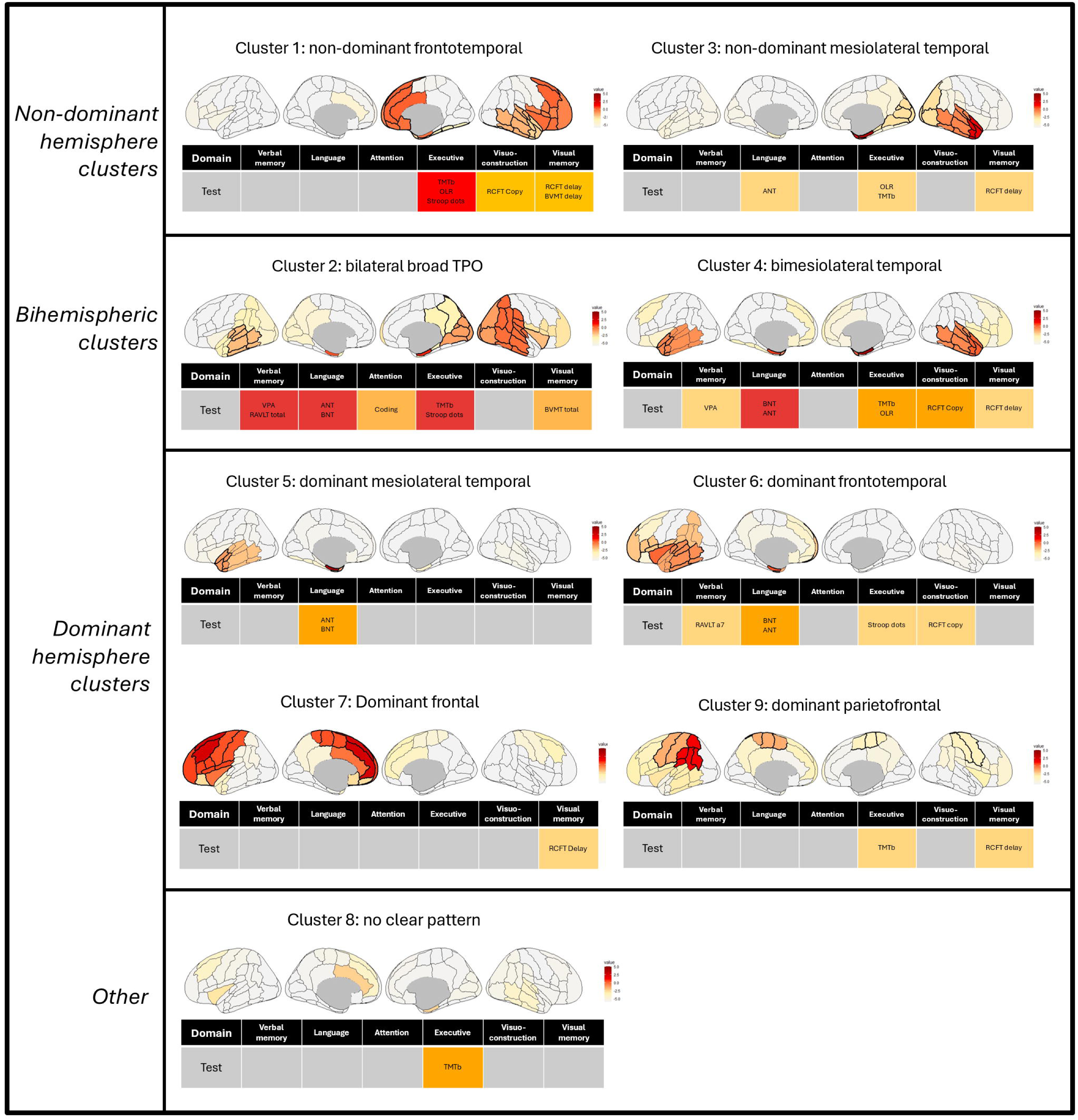
SEEG-Derived Epileptogenic Networks and Their Associated Cognitive Profiles. Cluster-specific patterns of epileptogenicity (top) and domain-level cognitive impairment (bottom). Brain schematics use the Harvard-Oxford cortical atlas. Bold outlines denote regions meeting both statistical (V-test > 1.65) and clinical (EzPz > –4) relevance thresholds. Colour gradients reflect epileptogenicity: orange-red (EzPz > –0.86, EZ) and grey-yellow (–6 to –0.86, PZ). Cognitive impairment is shown with a colour-coded heatmap: blue = none, yellow = mild, orange = moderate, red = severe.

### Epileptogenic network subtypes and their cognitive profiles

The following sections describe the epileptogenicity patterns and associated cognitive deficits for each network subtype. Demographic and clinical characteristics of the epileptic network clusters are summarised in Table 3.

### Non-dominant hemisphere clusters

Cluster 1 (n=4), ‘frontotemporal’: shows non-dominant frontal, insular, and temporal epileptogenicity. Severe executive dysfunction is accompanied by moderate impairments in visuoconstructional and visual memory measures.

Cluster 3 (n=4), ‘mesiolateral temporal’: is characterised by non-dominant mesiolateral temporal epileptogenicity with posterior quadrant extension and minor dominant temporal involvement. Mild impairments are observed in visual memory, executive function, and language.

### Bilateral clusters

Cluster 2 (n=3), ‘bilateral temporoparietooccipital’: shows bilateral posterior temporal epileptogenicity with parieto-occipital extension, predominantly in the non-dominant hemisphere. The network spans the temporo-parieto-occipital region. Severe impairments are observed in verbal memory, language, and executive function, with moderate deficits in attention and visual memory.

Cluster 4 (n=4), ‘bimesiolateral temporal’: epileptogenicity is highest in the bimesial temporal regions, with bilateral fronto-insular extension. Severe language impairment is accompanied by moderate deficits in executive function and visuoconstruction, and mild deficits in visual and verbal memory. This cluster also had the lowest TOPF = –1.5.

### Dominant hemisphere clusters

Cluster 5 (n=7), ‘mesiolateral temporal’: this cluster is characterised by dominant mesiolateral temporal epileptogenicity. Mild deficits are observed in language measures only. This cluster had a restricted mean network size (1.6 regions per patient); 6/7 patients were MRI-negative with one MRI-positive patient having hippocampal sclerosis.

Cluster 6 (n=4), ‘frontotemporal’: dominant frontal–insular–temporal epileptogenicity is observed, with minor extension into the dominant parietal region. Moderate language impairment is accompanied by mild deficits in verbal memory, executive function, and visuoconstruction.

Cluster 7 (n=4), ‘frontal’: epileptogenicity is restricted to the dominant frontal lobe. This cluster is associated with a mild deficit in visual memory only. This cluster had the highest TOPF.

Cluster 9 (n=6), ‘parietofrontal’: highest epileptogenicity is observed in the dominant parietal and posterior frontal regions, with additional involvement of the dominant temporal lobe. Broader bilateral frontoparietal extension is also present. Moderate impairments are seen in executive function and visual memory.

### Other

Cluster 8 (n=6): does not exhibit a clear pattern of epileptogenicity. An isolated moderate deficit is seen in executive function only. This cluster involved a greater number of regions across patients, but restricted size per patient (1.3 regions) reflecting a more heterogeneous group with more focal networks.

## Discussion

This proof-of-concept study explored the relationship between epileptic network organization and cognitive profiles. Using a composite SEEG-derived epileptogenicity metric (the EzPz score), we quantified regional epileptogenic burden in 42 SEEG patients. Dimensionality reduction and clustering identified nine epileptic network configurations that showed discernible patterns of cognitive impairment. These findings support a network-informed framework for interpreting cognitive dysfunction in epilepsy and illustrate the potential of this approach to inform diagnostic neuropsychology.

### Credibility of data-driven epileptic network subtypes

The clusters identified in this study should be interpreted as exploratory network configurations rather than definitive and stable epileptic subtypes given the modest sample size and variability of SEEG implantation locations.

The nine epileptic network configurations reflect recurring spatial configurations of epileptogenicity that are routinely encountered in clinical SEEG practice. These configurations were derived from clustering sublobar EzPz scores. Selected clusters are described below to highlight their clinical credibility.

Non-dominant and dominant frontotemporal epileptic networks were identified in clusters 1 and 6. These patterns align with the electroclinical entity of temporo-frontal epilepsy, a subtype of temporal-plus epilepsy^38,39^; studies utilizing EI provide intracranial evidence of this subtype.^40^

Clusters 3 and 5 reveal non-dominant and dominant mesiolateral temporal epileptic networks. The existence of mesiolateral temporal networks has also been demonstrated using coherence analysis^41^ and EI^40^; this subtype too is frequently encountered in clinical practice.

Clusters 2 and 9, both posteriorly organised, were characterised by multilobar epileptogenicity, which is characteristic of posterior epilepsies.^42^

Overall, the network configurations identified here are consistent with prior SEEG and biomarker-based descriptions. However, not all network subtypes were expected to resolve into nine clusters. For example, TLE has been subdivided into basal, polar, and posterior-superior variants^43,44^, and multiple prefrontal subtypes have been described^45^. These likely went unrepresented due to small subgroup sizes.

### Epileptic networks are associated with distinct and meaningful cognitive correlates

#### Pairwise relationships between brain regions and cognitive function

We first explored whether focal associations could be detected between epileptogenicity in discrete brain regions and cognitive impairments.

Exploratory pairwise analysis (Fig. 1) revealed significant negative correlations between sublobar epileptogenicity and domain-specific impairments. For example, language impairment correlated with dominant temporal epileptogenicity, and visual memory impairment with non-dominant mesial temporal involvement. These well-established structure–function relationships^9^ served as an internal plausibility check for the epileptogenicity metric and supported its use for exploratory network analyses.

Unexpected correlations also emerged—non-dominant frontal epileptogenicity was linked to visual memory impairment, and executive function appeared preserved despite dominant frontal involvement. Notably, verbal memory was not strongly associated with dominant mesial temporal epileptogenicity. These patterns demonstrate the limitations of pairwise comparisons, as such findings may reflect network interactions.

#### Epileptic network subtypes are associated with distinct cognitive profiles

We found that each of the nine epileptic network subtypes was associated with a unique cognitive signature (Fig. 3 and 4). Two important limitations are noted upfront. Firstly, the small sample size results in the exclusion of some network subtypes and thus prevents a review of associated cognitive patterns. Second, small cluster memberships preclude inferential statistics, so the ensuing discussion is necessarily qualitative.

Nevertheless, our proof-of-concept study suggests that the anatomical organization of epileptogenicity is meaningfully associated with cognitive impairment.

### Representative network–cognitive profile relationships

In cluster 1, the non-dominant frontotemporal network, severe deficits are seen in the executive domain and moderate deficits in visual memory and visuoconstructional domains. These impairments align with classical neuropsychological expectations, as FLE is consistently linked to deficits in executive function.^46^ The co-occurrence of visuoconstructional and visual memory impairments is plausible, reflecting the non-dominant hemisphere’s role in non-verbal functions.^47^ The visuoconstruction deficit may also reflect executive dysfunction, such as poor planning and organization, due to low construct specificity of the test.^48^

Cluster 6, the dominant frontotemporal network was associated with expected executive dysfunctions such as impaired response inhibition, as well as deficits in frontally mediated delayed verbal memory retrieval.^49^ Naming impairment was also present, consistent with dominant lateral temporal lobe involvement^50^, and may be further amplified by ventral prefrontal cortex engagement—a known language hub.^51^

Cluster 5, the dominant mesiolateral temporal network, is notable for the presence of naming impairment but without verbal memory deficits. Naming impairment reflects dominant lateral temporal involvement.^50^ However, preserved verbal memory is contrary to expectations for dominant mesial temporal involvement.^52^ Similarly, no significant association was found in the pairwise analysis. Six out of seven Cluster 5 patients were MRI-negative—a pattern common in modern SEEG cohorts—which may help explain the absence of verbal memory deficits. Classical expectations linking mesial temporal involvement to verbal memory impairment are largely based on earlier studies of patients with hippocampal sclerosis.^53^ While more recent work has shown that MRI-negative TLE can involve significant memory deficits^54^, this has not been stratified by network subtype. Our findings may suggest a distinct variant—MRI-negative dominant mesiolateral TLE—in which naming is disproportionately affected, challenging the assumption that mesial involvement necessarily predicts verbal memory impairment.

In contrast, Cluster 4—the bimesiolateral temporal network—showed verbal memory impairment, as well as deficits in language, visual memory, and visuoconstruction. This pattern aligns with expectations for bilateral temporal involvement. The broader cognitive profile may also reflect the larger network extent in this cluster, as indicated by a higher mean number of unique brain regions involved. Compared to Cluster 5, Cluster 4 demonstrates a more widespread, multidomain impairment profile.

Some unexpected associations were also observed. Cluster 7 (dominant frontal) lacked executive dysfunction and cluster 6 (dominant fronto-temporal) showed a mild visuoconstructional deficit. These findings may reflect underlying complex anatomical configurations, spanning multiple lobes with complex interactions, thereby deviating from syndromic-based expectations. However, they may also result from sample limitations, as unrepresented subtypes could exhibit alternative cognitive profiles. Nevertheless, a network-first approach may help uncover non-canonical cognitive patterns that are not predicted by traditional syndromic classifications Prior research supports a network-based view of cognitive dysfunction in epilepsy. Structural and functional imaging studies have shown that cortical thinning, white matter disruption, and altered connectivity extend beyond the EZ and correlate with cognitive deficits.^1,15,18,19^ Epileptic networks also exhibit abnormal synchrony and propagation in both ictal and interictal states^4,55,56^, and these alterations in network connectivity have been linked to impairments in memory, language, and executive function.^57^

Our findings extend this literature by demonstrating systematic relationships between sublobar epileptogenicity and cognitive impairment patterns. Specifically, we show that highly epileptogenic components of the broader propagation network—regions involved in early seizure spread and with a high burden of interictal discharges—may disrupt functional systems and contribute to cognitive deficits. By incorporating both the EZ and PZ, our approach captures dysfunctional elements of the network that may be missed in EZ-only analyses, but nonetheless meaningfully drive cognitive deficits.

Taken together, our results support the established role of diagnostic neuropsychology in presurgical evaluation—specifically, in generating hypotheses about the epileptic network to guide SEEG implantation and inform surgical decision-making. We propose that a network-informed framework may enhance this role, as the cognitive profiles it generated using a data-driven approach could support more precise and anatomically grounded hypotheses about network localization.

### Limitations

This study has several limitations. First, the modest sample size and subdivision into nine clusters reduced statistical power and precluded inferential statistics of cognitive profiles across clusters. While fewer clusters would have increased cluster membership, this approach was not supported by the data structure or clinical interpretability. The finer-grained clustering instead reflects the observed heterogeneity of epileptic networks seen in SEEG practice. Additionally, we did not control for clinical variables such as epilepsy onset age, duration, seizure burden, medication use, or epilepsy aetiology. Given the multidetermined nature of cognitive outcomes^58^, this is relevant in the context of small clusters. Given the modest cohort size and small cluster memberships, multivariable adjustment would have been statistically unstable and potentially misleading; therefore the analyses were intentionally descriptive.

Second, the EzPz score is a novel epileptogenicity metric. While grounded in validated methods of visually determined ictal onset, propagation^37^ and interictal spike rates, it does not incorporate additional quantitative or connectivity-based features that may further characterize epileptic networks. These include functional connectivity metrics, EI, cortical stimulation data, and time–frequency features such as LVFA or pre-ictal spiking. Although the EzPz score has not yet undergone external validation and its generalisability is uncertain, internal validation across seizure-free and non–seizure-free cohorts has been encouraging (see Supplementary Fig. 1).

Third, SEEG sampling inherently limits complete network characterisation, which may bias interpretation of network–cognition relationships. This limitation is common to SEEG-based biomarkers and underscores the need for multimodal integration with neuroimaging.

### Future directions

Future studies would benefit from larger, multicentre cohorts to improve statistical power and allow for inferential comparisons of cognitive profiles across clusters. Larger cohorts will also reduce the influence of confounding variables on neuropsychological outcomes. Finer clustering solutions could also be achieved, uncovering clinically meaningful network subtypes not captured here.

Larger datasets could support multivariate techniques to formally test the distinctiveness of neuropsychological patterns. Given the relative infrequency of SEEG, international collaborations will be critical to achieve these aims.

Future work should aim to further validate the EzPz score and examine its clinical utility. Importantly, the overall framework is flexible and can also utilize established and emerging biomarkers such as EI, HFO and connectivity metrics.

### Conclusion

This study presents a SEEG-based, data-driven framework for linking epileptic networks to patterns of neuropsychological impairment. By moving beyond syndrome-based models toward a network-level perspective, this approach provides a framework for conceptualising cognitive dysfunction in epilepsy. With further validation in larger cohorts, network-specific cognitive profiles may help generate presurgical hypotheses regarding network localisation and lateralisation, potentially informing electrode implantation strategies and surgical planning.

## Data availability

The data that support the findings of this study are available from the corresponding author, upon reasonable request

## Author Contributions

P.S. conceived and designed the study, performed data acquisition and statistical analyses, interpreted the data, and drafted the manuscript.

A.N. contributed to study conception and design and critically revised the manuscript.

E.C. contributed to data acquisition and manuscript revision.

T.W. contributed to statistical analyses.

G.R. and M.H. contributed to manuscript drafting and revision.

All remaining authors contributed to critical revision of the manuscript for important intellectual content.

---

Supplementary Figure 1: **Correlation matrix with annotated coefficients between regional epileptogenicity and neuropsychological performance**

Same correlation matrix as Figure 1, with annotated Pearson’s r values. Negative coefficients indicate that greater epileptogenicity is associated with poorer performance on neuropsychological testing.

Supplementary Figure 2. **EzPz scores distinguish thermocoagulated contacts in seizure-free and full patient cohorts.**

**A**: Distribution of EzPz scores in thermocoagulated (RF-THC) versus non-thermocoagulated contacts among seizure-free patients (N = 13). EzPz scores were significantly higher in RF-THC contacts (*p* < 0.001), supporting an association between the score and contacts targeted for ablation (i.e. part of the EZ).

**B**: Distribution of EzPz scores across all contacts in the full patient cohort (N = 52), including both seizure-free and non-seizure-free patients. Scores remained significantly elevated in RF-THC contacts (*p* < 0.001), demonstrating generalizability of the score.

## References

1. Hermann B, Loring DW, Wilson S. Paradigm Shifts in the Neuropsychology of Epilepsy. Journal of the International Neuropsychological Society. 2017;23(9-10):791–805. doi:10.1017/s1355617717000650

2. Baxendale S, Thompson P. The new approach to epilepsy classification: Cognition and behavior in adult epilepsy syndromes. Epilepsy & Behavior. 2016;64:253–256. doi:10.1016/j.yebeh.2016.09.003

3. Spencer SS. Neural networks in human epilepsy: evidence of and implications for treatment. Epilepsia. Mar 2002;43(3):219–27. doi:10.1046/j.1528-1157.2002.26901.x

4. Bartolomei F, Lagarde S, Wendling F, et al. Defining epileptogenic networks: Contribution of SEEG and signal analysis. Epilepsia. Jul 2017;58(7):1131–1147. doi:10.1111/epi.13791

5. Bressler SL, Menon V. Large-scale brain networks in cognition: emerging methods and principles. Trends Cogn Sci. Jun 2010;14(6):277–90. doi:10.1016/j.tics.2010.04.004

6. Wilson SJ, Baxendale S. The new approach to classification: rethinking cognition and behavior in epilepsy. Epilepsy Behav. Dec 2014;41:307–10. doi:10.1016/j.yebeh.2014.09.011

7. Kramer MA, Cash SS. Epilepsy as a Disorder of Cortical Network Organization. The Neuroscientist. 2012-08-01 2012;18(4):360–372. doi:10.1177/1073858411422754

8. Ryvlin P. SEEG in 2025: progress and pending challenges in stereotaxy methods, biomarkers and radiofrequency thermocoagulation. Curr Opin Neurol. Apr 1 2025;38(2):111–120. doi:10.1097/WCO.0000000000001351

9. Jones-Gotman M, Smith ML, Risse GL, et al. The contribution of neuropsychology to diagnostic assessment in epilepsy. Epilepsy Behav. May 2010;18(1-2):3–12. doi:10.1016/j.yebeh.2010.02.019

10. Hermann B, Seidenberg M, Lee EJ, Chan F, Rutecki P. Cognitive phenotypes in temporal lobe epilepsy. J Int Neuropsychol Soc. Jan 2007;13(1):12–20. doi:10.1017/S135561770707004X

11. Arrotta K, Reyes A, Kaestner E, et al. Cognitive phenotypes in frontal lobe epilepsy. Epilepsia. 2022;doi:10.1111/epi.17260

12. Reyes A, Kaestner E, Ferguson L, et al. Cognitive phenotypes in temporal lobe epilepsy utilizing data- and clinically driven approaches: Moving toward a new taxonomy. Epilepsia. Jun 2020;61(6):1211–1220. doi:10.1111/epi.16528

13. Dabbs K, Jones J, Seidenberg M, Hermann B. Neuroanatomical correlates of cognitive phenotypes in temporal lobe epilepsy. Epilepsy Behav. Aug 2009;15(4):445–51. doi:10.1016/j.yebeh.2009.05.012

14. Hermann BP, Lin JJ, Jones JE, Seidenberg M. The emerging architecture of neuropsychological impairment in epilepsy. Neurol Clin. Nov 2009;27(4):881–907. doi:10.1016/j.ncl.2009.08.001

15. Reyes A, Kaestner E, Bahrami N, et al. Cognitive phenotypes in temporal lobe epilepsy are associated with distinct patterns of white matter network abnormalities. Neurology. Apr 23 2019;92(17):e1957–e1968. doi:10.1212/WNL.0000000000007370

16. Norman M, Wilson SJ, Baxendale S, et al. Addressing neuropsychological diagnostics in adults with epilepsy: Introducing the International Classification of Cognitive Disorders in Epilepsy: The IC CODE Initiative. Epilepsia Open. Jun 2021;6(2):266–275. doi:10.1002/epi4.12478

17. Rosenow F, Lüders H. Presurgical evaluation of epilepsy. Brain. 2001;124(9):1683–1700. doi:10.1093/brain/124.9.1683

18. Bruce H, Michael S, Brian B, et al. Extratemporal quantitative MR volumetrics and neuropsychological status in temporal lobe epilepsy. Journal of the International Neuropsychological Society. 2003;9(3):353–362. doi:10.1017/S1355617703910113

19. Baxendale SA, Sisodiya SM, Thompson PJ, et al. Disproportion in the distribution of gray and white matter. Neurology. 1999;52(2):248. doi:10.1212/WNL.52.2.248

20. Marla J H, William T S. Auditory and visual naming tests: Normative and patient data for accuracy, response time, and tip-of-the-tongue. Journal of the International Neuropsychological Society. 2003;9(3):479–489. doi:10.1017/S135561770393013X

21. Wechsler D. Wechsler memory scale-revised. Psychological Corporation. 1987;

22. Benedict RH, Schretlen D, Groninger L, Dobraski M, Shpritz B. Revision of the Brief Visuospatial Memory Test: Studies of normal performance, reliability, and validity. Psychological assessment. 1996;8(2):145.

23. Wechsler D. Manual for the Wechsler adult intelligence scale. 1955;

24. Reitan R. The Halstead-Reitan neuropsychological test battery: therapy and clinical interpretation. (No Title). 1985;

25. Esther Strauss EMSS, Otfried Spreen. A Compendium of Neuropsychological Tests_ Administration, Norms, and Commentary. 2006;

26. Kaplan E, Goodglass H, Weintraub S. Boston naming test. The Clinical Neuropsychologist. 2001;

27. Spreen O, Strauss E. A compendium of neuropsychological tests: Administration, norms, and commentary. Oxford University Press; 1998.

28. Rikir E, Koessler L, Gavaret M, et al. Electrical source imaging in cortical malformation-related epilepsy: a prospective EEG-SEEG concordance study. Epilepsia. Jun 2014;55(6):918–32. doi:10.1111/epi.12591

29. Roehri N, Pizzo F, Lagarde S, et al. High-frequency oscillations are not better biomarkers of epileptogenic tissues than spikes. Ann Neurol. Jan 2018;83(1):84–97. doi:10.1002/ana.25124

30. Bartolomei F, Trebuchon A, Bonini F, et al. What is the concordance between the seizure onset zone and the irritative zone? A SEEG quantified study. Clin Neurophysiol. Feb 2016;127(2):1157–1162. doi:10.1016/j.clinph.2015.10.029

31. Shi W, Shaw D, Walsh KG, et al. Spike ripples localize the epileptogenic zone best: an international intracranial study. Brain. Jul 5 2024;147(7):2496–2506. doi:10.1093/brain/awae037

32. Thomas J, Kahane P, Abdallah C, et al. A Subpopulation of Spikes Predicts Successful Epilepsy Surgery Outcome. Ann Neurol. Mar 2023;93(3):522–535. doi:10.1002/ana.26548

33. Grinenko O, Li J, Mosher JC, et al. A fingerprint of the epileptogenic zone in human epilepsies. Brain. Jan 1 2018;141(1):117–131. doi:10.1093/brain/awx306

34. Di Giacomo R, Burini A, Chiarello D, et al. Ictal fast activity chirps as markers of the epileptogenic zone. Epilepsia. Jun 2024;65(6):e97–e103. doi:10.1111/epi.17995

35. Bartolomei F, Chauvel P, Wendling F. Epileptogenicity of brain structures in human temporal lobe epilepsy: a quantified study from intracerebral EEG. Brain. Jul 2008;131(Pt 7):1818–30. doi:10.1093/brain/awn111

36. Lagarde S, Buzori S, Trebuchon A, et al. The repertoire of seizure onset patterns in human focal epilepsies: Determinants and prognostic values. Epilepsia. Jan 2019;60(1):85–95. doi:10.1111/epi.14604

37. Bonini F, McGonigal A, Trebuchon A, et al. Frontal lobe seizures: from clinical semiology to localization. Epilepsia. Feb 2014;55(2):264–77. doi:10.1111/epi.12490

38. Kahane P, Barba C, Rheims S, Job-Chapron AS, Minotti L, Ryvlin P. The concept of temporal ‘plus’ epilepsy. Revue neurologique. 2015;171(3):267–272. doi:10.1016/j.neurol.2015.01.562

39. Barba C, Barbati G, Minotti L, Hoffmann D, Kahane P. Ictal clinical and scalp-EEG findings differentiating temporal lobe epilepsies from temporal ‘plus’ epilepsies. Brain. 2007;130(7):1957–1967. doi:10.1093/brain/awm108

40. Bartolomei F, Cosandier-Rimele D, Mcgonigal A, et al. From mesial temporal lobe to temporoperisylvian seizures: A quantified study of temporal lobe seizure networks. Epilepsia. 2010-10-01 2010;51(10):2147-2158. doi:10.1111/j.1528-1167.2010.02690.x

41. Bartolomei F, Wendling F, Vignal J-P, et al. Seizures of temporal lobe epilepsy: identification of subtypes by coherence analysis using stereo-electro-encephalography. Clinical Neurophysiology. 1999/10/01/ 1999;110(10):1741-1754. 10.1016/S1388-2457(99)00107-8

42. Marchi A, Bonini F, Lagarde S, et al. Occipital and occipital “plus” epilepsies: A study of involved epileptogenic networks through SEEG quantification. Epilepsy Behav. Sep 2016;62:104–14. doi:10.1016/j.yebeh.2016.06.014

43. Hadidane S, Lagarde S, Medina-Villalon S, et al. Basal temporal lobe epilepsy: SEEG electroclinical characteristics. Epilepsy Res. Mar 2023;191:107090. doi:10.1016/j.eplepsyres.2023.107090

44. Chabardes S, Kahane P, Minotti L, et al. The temporopolar cortex plays a pivotal role in temporal lobe seizures. Brain. Aug 2005;128(Pt 8):1818–31. doi:10.1093/brain/awh512

45. Machado S, Bonini F, Mcgonigal A, et al. Prefrontal seizure classification based on stereo-EEG quantification and automatic clustering. Epilepsy & Behavior. 2020-11-01 2020;112:107436. doi:10.1016/j.yebeh.2020.107436

46. Patrikelis P, Angelakis E, Gatzonis S. Neurocognitive and behavioral functioning in frontal lobe epilepsy: A review. Epilepsy & Behavior. 2009;14(1):19–26. doi:10.1016/j.yebeh.2008.09.013

47. Jones-Gotman M, Milner B. Right temporal-lobe contribution to image-mediated verbal learning. Neuropsychologia. 1978/01/01/ 1978;16(1):61–71. 10.1016/0028-3932(78)90043-X

48. Diaz-Orueta U, Rogers BM, Blanco-Campal A, Burke T. The challenge of neuropsychological assessment of visual/visuo-spatial memory: A critical, historical review, and lessons for the present and future. Frontiers in Psychology. 2022;13:962025.

49. Loring DW, Strauss E, Hermann BP, et al. Differential neuropsychological test sensitivity to left temporal lobe epilepsy. Journal of the International Neuropsychological Society. 2008;14(3):394–400.

50. Hermann BP, Seidenberg M, Haltiner A, Wyler AR. Adequacy of Language Function and Verbal Memory Performance in Unilateral Temporal Lobe Epilepsy. Cortex. 1992;28(3):423–433. doi:10.1016/s0010-9452(13)80152-9

51. Arrotta K, Swanson SJ, Janecek JK, et al. Application of the International Classification of Cognitive Disorders in Epilepsy (IC-CoDE) to frontal lobe epilepsy using multicenter data. Epilepsy & Behavior. 2023/11/01/ 2023;148:109471. 10.1016/j.yebeh.2023.109471

52. Hermann BP, Seidenberg M, Schoenfeld J, Davies K. Neuropsychological characteristics of the syndrome of mesial temporal lobe epilepsy. Arch Neurol. Apr 1997;54(4):369–76. doi:10.1001/archneur.1997.00550160019010

53. Baxendale SA, van Paesschen W, Thompson PJ, et al. The Relationship Between Quantitative MRI and Neuropsychological Functioning in Temporal Lobe Epilepsy. 10.1111/j.1528-1157.1998.tb01353.x. Epilepsia. 1998/02/01 1998;39(2):158-166. 10.1111/j.1528-1157.1998.tb01353.x

54. Rayner G, Tailby C, Jackson G, Wilson S. Looking beyond lesions for causes of neuropsychological impairment in epilepsy. Neurology. Feb 12 2019;92(7):e680–e689. doi:10.1212/wnl.0000000000006905

55. Centeno M, Carmichael DW. Network Connectivity in Epilepsy: Resting State fMRI and EEG-fMRI Contributions. Front Neurol. 2014;5:93. doi:10.3389/fneur.2014.00093

56. Bettus G, Ranjeva JP, Wendling F, et al. Interictal functional connectivity of human epileptic networks assessed by intracerebral EEG and BOLD signal fluctuations. PLoS One. 2011;6(5):e20071. doi:10.1371/journal.pone.0020071

57. Englot DJ, Konrad PE, Morgan VL. Regional and global connectivity disturbances in focal epilepsy, related neurocognitive sequelae, and potential mechanistic underpinnings. Epilepsia. 2016-10-01 2016;57(10):1546-1557. doi:10.1111/epi.13510

58. Baxendale S, Thompson P. Beyond localization: The role of traditional neuropsychological tests in an age of imaging. Epilepsia. 2010-11-01 2010;51(11):2225–2230. doi:10.1111/j.1528-1167.2010.02710.x

