## Supplementary figures and images for "A Data-Driven Approach for Linking Epileptic Networks and Cognitive Profiles Using Stereo-EEG"

### Supplemental Figure 1

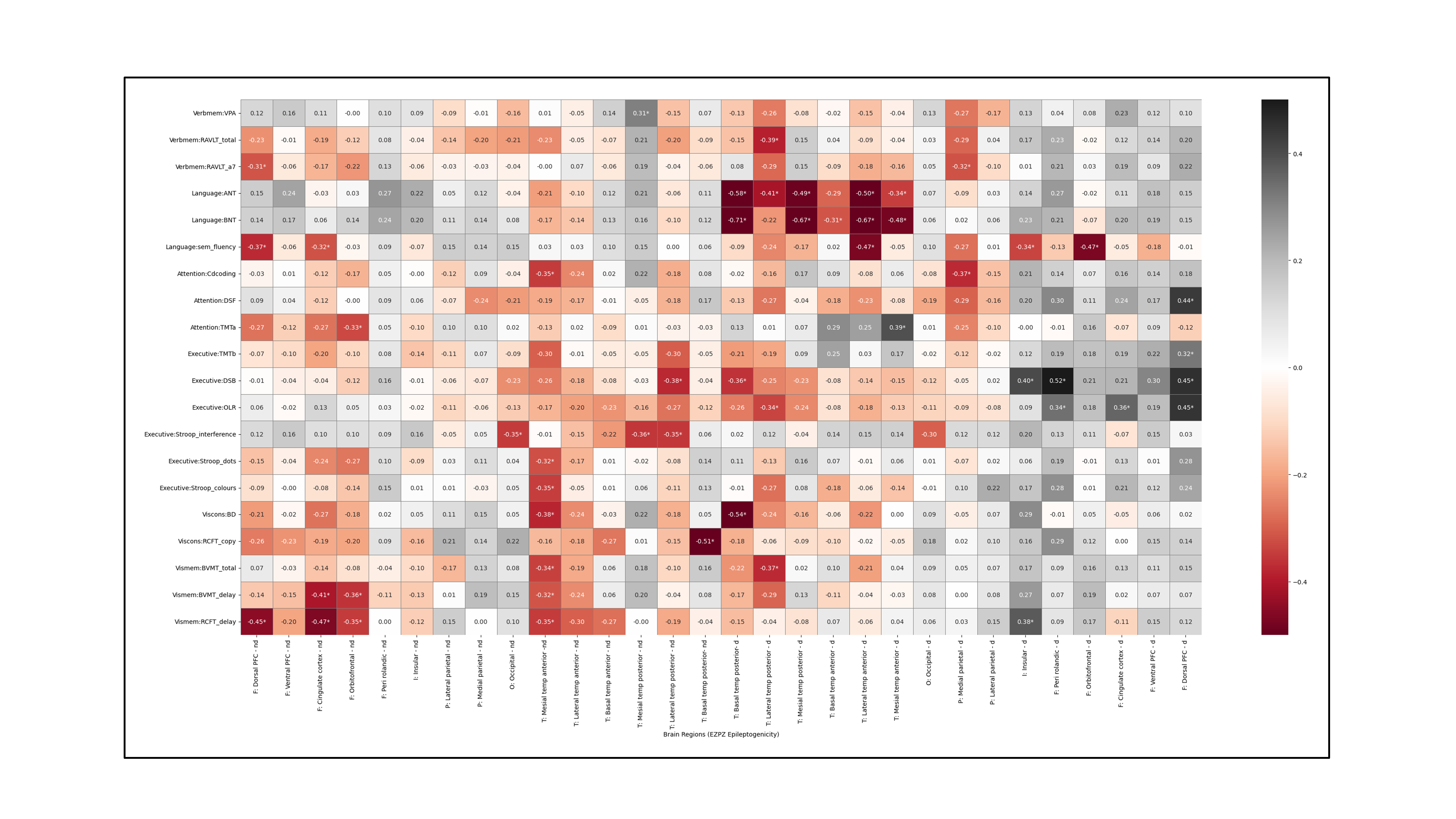

### Supplemental Figure 2

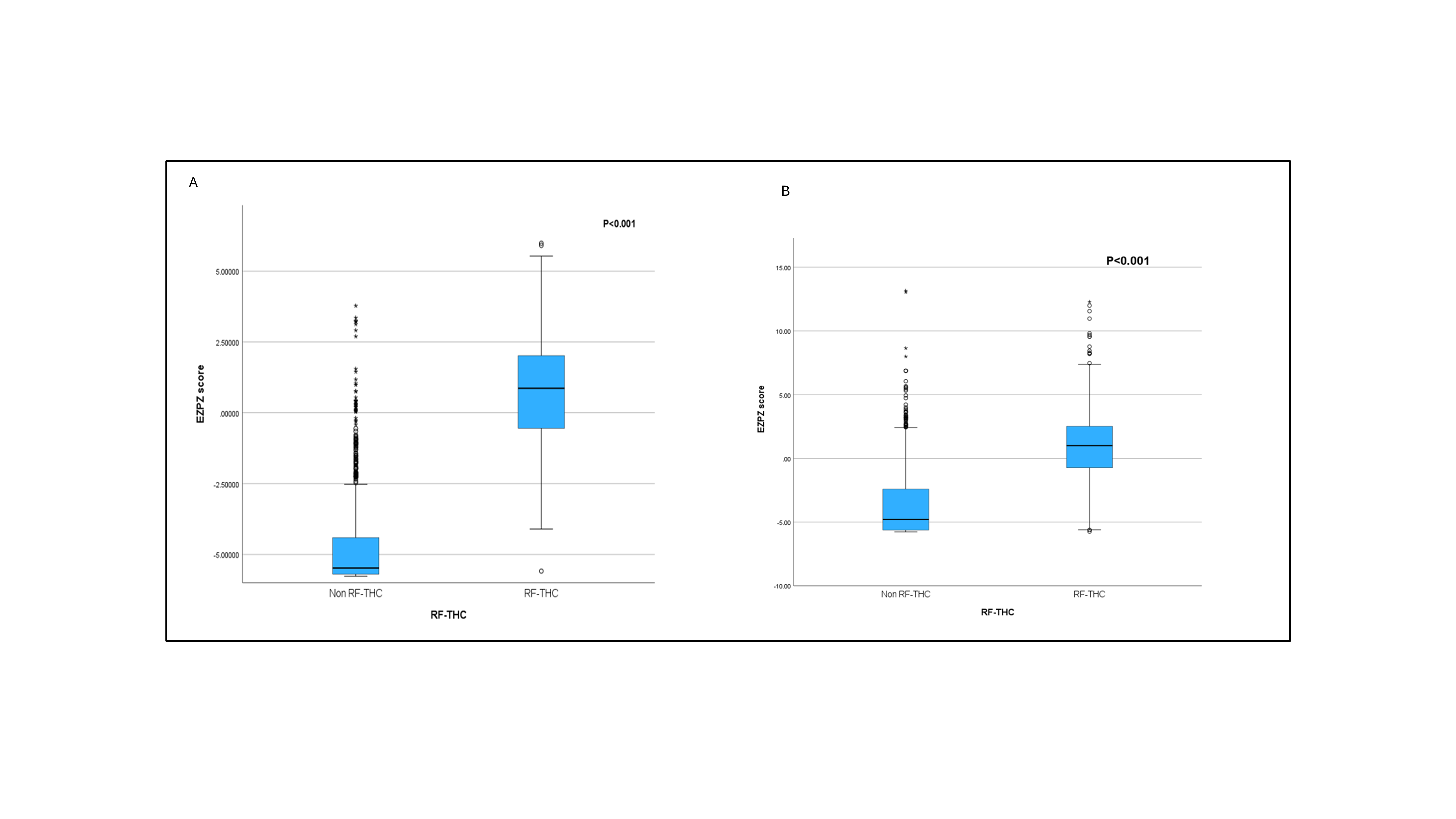
