## Supplemental Table 1 for "A Data-Driven Approach for Linking Epileptic Networks and Cognitive Profiles Using Stereo-EEG"

### Supplementary

**Supplementary Table1: Predefined brain regions used for sublobar contact categorisation**

| Brain Region[a] |
|---|
| Premotor–motor cortex |
| Dorsal prefrontal cortex |
| Ventral prefrontal cortex |
| Medial frontal cortex |
| Orbitofrontal cortex |
| Insular cortex |
| Medial anterior temporal cortex |
| Lateral anterior temporal cortex |
| Basal temporal cortex |
| Lateral posterior temporal cortex |
| Medial parietal cortex |
| Lateral parietal cortex |
| Occipital cortex |

[a]Each region was classified separately for the dominant and non-dominant hemispheres, yielding a total of 26 sublobar labels per patient.

#### Correlation matrix with annotated coefficients between regional epileptogenicity and neuropsychological performance

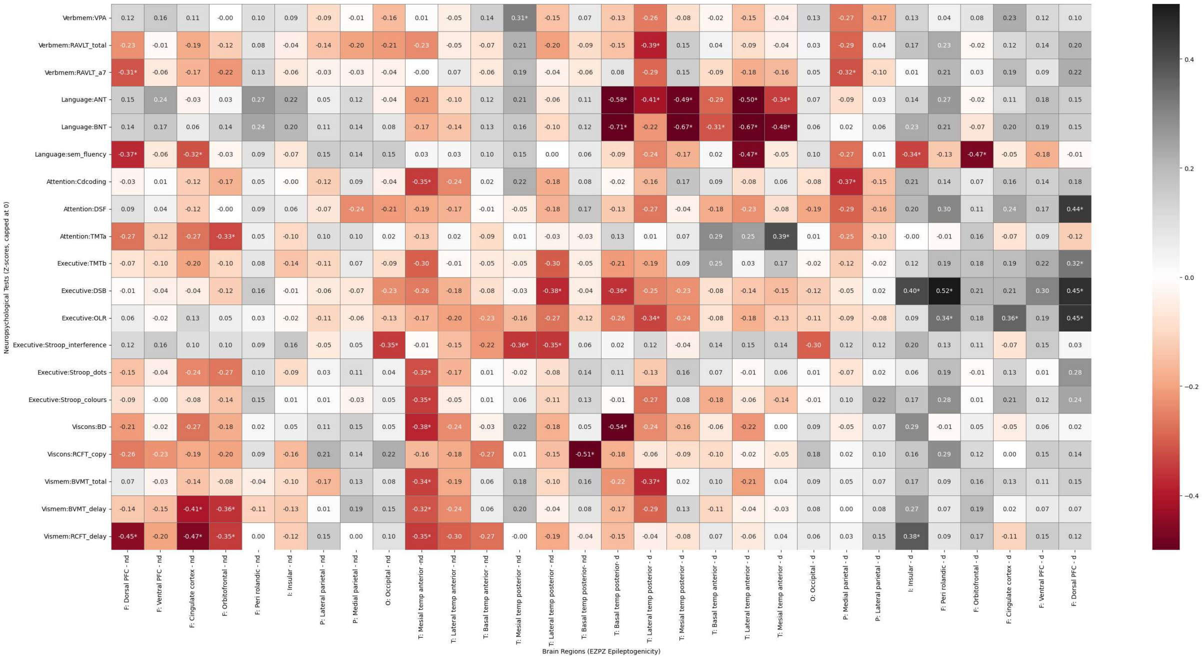


Supplementary Figure 1: **Correlation matrix with annotated coefficients between regional epileptogenicity and neuropsychological performance**

Same correlation matrix as Figure 1, with annotated Pearson's r values. Negative coefficients indicate that greater epileptogenicity is associated with poorer performance on neuropsychological testing.

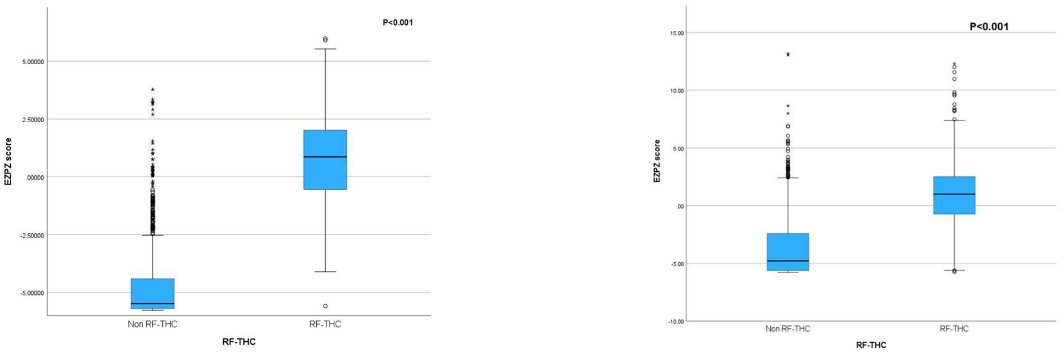


**Supplementary Figure 2: EzPz Scores Distinguish Thermocoagulated Contacts in Seizure-Free and Full Cohorts**

**A** Distribution of EzPz scores in thermocoagulated (RF-THC) versus non-thermocoagulated contacts among seizure-free patients (N = 13). EzPz scores were significantly higher in RF-THC contacts ($p < 0.001$), supporting the association between the score and contacts targeted for ablation ( as part of EZ).

**B** Distribution of EzPz scores across all contacts in the full patient cohort (N = 52). This includes both seizure-free and non-seizure-free patients. Scores remain significantly elevated in RF-THC contacts ($p < 0.001$), demonstrating generalizability of the score.
